# Impact of the COVID-19 pandemic on developmental care practices for infants born preterm

**DOI:** 10.1101/2020.11.25.20238956

**Authors:** Melissa Scala, Virginia A. Marchman, Edith Brignoni-Pérez, Maya Chan Morales, Katherine E. Travis

## Abstract

**Objectives:** To assess the impact of the COVID-19 pandemic on rates of hospital visitation and rates and durations of developmental care practices for infants born preterm delivered by both families and clinical staff.

**Methods:** We analyzed electronic medical record data from infants born at less than 32 weeks gestational age (GA) cared for in the Lucile Packard Children’s Hospital neonatal intensive care unit (NICU) in a COVID-19-affected period (March 8, 2020 to May 31, 2020) and the analogous period in 2019. Our final sample consisted of 52 infants (*n*=27, 2019 cohort; *n*=25, 2020 cohort). Rates of family visitation and of family- and clinical staff-delivered developmental care were compared across cohorts, adjusting for GA at start of study period.

**Results:** Results indicated that families of infants in the 2020 cohort visited less frequently (47% of available days) than those in the 2019 cohort (97%; *p*=0.001). Infants received developmental care activities less frequently in the 2020 cohort (3.51 vs. 4.72 activities per day; *p*=0.04), with a lower number of minutes per day (99.91 vs. 145.14; *p*=0.04) and a shorter duration per instance (23.41 vs. 29.65; *p*=0.03). Similar reductions occurred in both family- and staff-delivered developmental care activities.

**Conclusions:** The COVID-19 pandemic has negatively impacted family visitation and preterm infant developmental care practices in the NICU, both experiences associated with positive health benefits. Hospitals should create programs to improve family visitation and engagement, while also increasing staff-delivered developmental care. Careful attention should be paid to long-term follow up of preterm infants and families.

## Introduction

Each year, approximately 380,000 infants in the United States and 15 million worldwide are born preterm (before 37 weeks of gestation)^1^. Preterm children, especially those born very preterm (<32 weeks gestational age, GA) are at high risk for adverse health and neurodevelopmental outcomes that persist far beyond the newborn period^2^. Over the last 20 years, advances in perinatal developmental care practices, such as positive touch, skin-to-skin holding (kangaroo care), swaddled holding, massage, music therapy, and reading, have led to substantial improvements in health and neurodevelopmental outcomes of preterm children, including reduced rates of infections, and improved cardiovascular health, growth, and behavioral organization^3^. Studies have also shown that developmental care practices reduce parent and infant stress and improve parent-infant bonding and rates of breastfeeding^4^. Due to the COVID-19 pandemic, however, there is substantial concern that preterm infants may be deprived of access to developmental care activities and may therefore be more at-risk for poor health and long-term neurodevelopmental outcomes than preterm infants born prior to the pandemic^5,6^.

As a consequence of the COVID-19 pandemic, healthcare centers, including our center, instituted restricted visitation policies^7^, guided by recommendations from national experts at the Centers for Disease Control and Prevention, similar to many facilities globally^8^. These policies, combined with other factors that may limit families’ ability or willingness to visit their preterm infant in the neonatal intensive care unit (NICU) (e.g., concerns about disease transmission and/or limited access to childcare for siblings), may limit preterm infants’ access to developmental care activities, thereby, reducing opportunities for associated health benefits. In the present study, we sought to assess the impact of the COVID-19 pandemic on rates of family visitation and rates and durations of developmental care practices on infants born preterm delivered by both families and clinical staff. We hypothesized that families would have reduced rates of visitation and less engagement in developmental care activities in the COVID-19 period than in a similar period the year prior. We also hypothesized that clinical staff would deliver reduced amounts of developmental care as compared to those in previous period due to psychological aspects of physical distancing under the COVID-19 pandemic, but that their reduction in activities would be less than that of families, whose physical presence in the NICU was impacted.

## Methods

### Participants

Participants were infants (*N*=52; 20 females, 32 males) born very preterm (< 32 GA). Based on information in the Electronic Medical Records (EMR), all infants were either born or transferred for hospitalization in the NICU at Lucile Packard Children’s Hospital (LPCH) within one week of their birth and stayed for at least seven days during the study period in 2019 (March 8 - May 31, 2019 cohort, *n* = 27) and 2020 (March 8 - May 31, 2020 cohort, *n* = 25). An additional 19 infants were excluded from analyses because the number of days during the study period was less than seven days (*n*=1), the infant was admitted less than one week after birth (*n*=10), or both (*n* = 8). These exclusions allowed for similar postmenstrual age (PMA) between cohorts, with similar expected patterns of developmental care activity, and an adequate representative period of hospitalization in which developmental care activities could occur.

Table 1 shows that infants were primarily from non-White families and about half of the families in both cohorts used public, rather than private, health insurance. The infants were about 28 weeks GA, on average, with no difference between cohorts. Mean PMA at first day within the study period was significantly older for infants in the 2019 cohort than those in the 2020 cohort. Therefore, PMA at study period start is a covariate in all analyses. Number of days during the study period was not significantly different between cohorts, ranging from a minimum of one week to the complete study period in both groups (2019: 7 – 84, 2020: 9 – 84).

**Table 1.** Demographic and clinical factors of infants followed during study period in 2019 and 2020 (*N*=52).

| | Cohort | | $t$ or $\chi^2$ | $p$ |
| --- | --- | --- | --- | --- |
|  | 2019 | 2020 |  |  |
| $N$ | 27 | 25 | -- | -- |
| Female: $n$ (%) | 10 (37.0) | 10 (40.0) | 0.05 | 0.53 |
| White: $n$ (%) | 4 (14.8) | 1 (4.0) | 1.75 | 0.35 |
| Public Insurance: $n$ (%) | 13 (48.2) | 12 (48.0) | 0.01 | 0.99 |
| GA at birth (weeks)<sup>a</sup> | 28.6 (2.3) | 28.3 (2.2) | 0.56 | 0.58 |
| PMA<sup>b</sup> | 222.3 (28.3) | 208.0 (18.9) | 2.13 | 0.04* |
| Days in NICU<sup>c</sup> | 41.3 (25.7) | 50.0 (22.4) | 1.29 | 0.20 |
| Apgar 1 min<sup>d</sup> | 5.5 (2.1) | 4.9 (2.4) | 0.93 | 0.36 |
| Apgar 5 min<sup>d</sup> | 8.0 (2.3) | 7.6 (1.6) | 0.67 | 0.51 |
| NEC<sup>e</sup> | 4 (14.8) | 2 (8.0) | 0.59 | 0.67 |
| BPD<sup>f</sup> | 5 (18.5) | 3 (12.0) | 0.42 | 0.71 |
| IVH<sup>g</sup> | 5 (18.5) | 6 (24.0) | 0.23 | 0.74 |

\* $p < .05$
<sup>a</sup>Gestational age (GA) in weeks at birth.
<sup>b</sup>Post-menstrual age (PMA) at start of NICU hospitalization during study period.
<sup>c</sup>Number of days of NICU hospitalization during study period.
<sup>d</sup>Apgar score (10 max) taken at 1 min and 5 mins after birth.
<sup>e</sup>Necrotizing Enterocolitis (NEC)
<sup>f</sup>Bronchopulmonary Dysplasia (BPD)
<sup>g</sup>Intraventricular Hemorrhage (IVH)

To determine comparability in the health status of the infants in the 2019 versus the 2020 cohorts, several clinical variables were accessed from the EMR. As shown in Table 1, Apgar scores recorded at 1 minute and 5 minutes post-birth were not significantly different between cohorts. In addition, the incidence of Necrotizing Enterocolitis, Bronchopulmonary Dysplasia, and Intraventricular Hemorrhage were also similar across the two cohorts.

### Procedure and Measures

#### Study Period

Visitation and developmental care (DC) activities were retrospectively analyzed based on the EMR over two periods, during the COVID-19 pandemic (March 8 - May 31, 2020) and at an analogous period prior to the pandemic (March 8 - May 31, 2019). This study period was selected because March 8, 2020 was the first day on which pandemic-related visitation policies were implemented at LPCH and those policies were partially relaxed on June 1, 2020. Prior to March 8, 2020, parents and family members were permitted to visit at any time of the day, except during nursing sign out (7:00-7:30 a.m./p.m.). After March 8, 2020, state and county guidelines resulting from the COVID-19 pandemic required that LPCH visitation policies change. On that date, visitation was limited to parents only. On March 30, 2020, only one parent was allowed to visit per infant for the entire hospital stay. By May 7, 2020, parents could alternate days visiting but could not come together. By June 2020, both parents could again visit their infant but no other family members. In addition to these changes to visitation policies, physical distancing protocols for clinical staff were in effect starting March 8, 2020 with some members working remotely, reducing on-site clinical support. To offset hospital revenue loss, clinical staff took paid time off, further reducing developmental team and social work staffing. These changes did not impact bedside nursing ratios but reduced on-site family support services. No restrictions to DC activities occurred during either period. Family members and clinical staff were required to perform hand hygiene and to avoid the NICU with signs of illness (both 2019 and 2020 cohorts) and, additionally, to universally wear a mask (2020 Cohort).

Routine charting of all visitation and DC activities in the EMR was instituted on May 1, 2018 and therefore was well-established by the onset of the pre-pandemic study period in 2019. Visitation policy changes implemented due to the COVID-19 pandemic did not directly affect protocols regarding the charting of visitation and DC activities and therefore procedures required of clinical staff were identical for the 2019 and 2020 cohorts.

#### Family Visitation

To determine family visitation rates, each instance of family visitation was determined from the EMR. Visitation was assumed to occur if the clinical staff charted a visitation event and/or if any family member engaged in DC at bedside. Family visitation rate was the proportion of days with at least one instance of family visitation out of all days during the study period.

#### Developmental Care

As part of routine practice, clinical staff charted the occurrence of each instance of DC, the approximate duration, and who was involved, for seven types of DC activities: Kangaroo Care, Holding, Touch, Massage, Music, Talking, and Singing. Instances of DC were collapsed across type, given the differences in the PMA of the infants and hence which specific types of DC activities might be more or less likely to occur. However, all parents at LPCH are taught a range of DC activities appropriate for their infant so that all infants in this sample were offered some type of DC during the study period. *Frequency* was defined as the number of instances of DC out of number of days during study period, capturing the rate of DC per day normalized for each infant’s length of stay during the study period. *Amount* was the sum of all minutes of DC out of number of days during the study period, capturing the minutes per day of DC. *Duration* was the mean number of minutes per DC activity, calculated as the sum of all DC minutes out of the number of DC instances. Each measure was sub-categorized by who was involved in the activity to derive frequency, amount, and duration of DC involving *Family* or *Clinical Staff*.

#### Analytic Strategy

We first present descriptive statistics for the frequency, amount, and duration of DC by cohort (2019 vs. 2020). To explore cohort differences in visitation rates and measures of the frequency, amount, and duration of DC activities, we conducted univariate analyses of co-variance (ANCOVA) with cohort (2019 vs. 2020) as the between-group factor, controlling for PMA at start of study period. To explore differences in DC activities as a function of who delivered the care, we conducted mixed ANCOVAs on each measure with DC delivery source (Family vs. Clinical Staff) as the within-group factor and cohort (2019 vs. 2020) as the between-group factor, again controlling for PMA at start of study period. Effect sizes were estimated using Cohen’s *d*, expressed in SD units. Significance levels were set at *p* < 0.05 for all analyses. The protocols for this study were approved by the Stanford University Institutional Review Board. Participants were not required to provide consent, because this study was based on a retrospective chart review.

## Results

Table 2 shows that, on average, families in the 2019 cohort visited the hospital more than 95% of the days their infant was in the hospital. During 2020, rates of visitation dropped significantly to less than half of all possible days. In addition, infants in the 2019 cohort experienced approximately 4.8 activities per day, compared to approximately 3.5 DC activities per day in the 2020 cohort, on average, a reduction of approximately 1.2 DC activities per day (*d* = .55). Moreover, infants in the 2019 cohort experienced approximately 50 more minutes of DC per day than infants in the 2020 cohort (*d* = .52), with each DC instance lasting about 6 minutes longer in 2019, on average, than in 2020 (*d* = .60).

**Table 2.** Estimated marginal means (SD) and group comparisons controlling for PMA at start of study period for rates of visitation and frequency, amount, and duration of developmental care (DC) by cohort (2019, 2020) (*N*=52).

| | Cohort | | $F(2,52)$ | $p$ |
| --- | --- | --- | --- | --- |
|  | 2019 | 2020 |  |  |
| Family Visitation<sup>a</sup> | .97 (.22) | .47 (.24) | 59.13 | .001** |
| Frequency<sup>b</sup> | 4.72 (1.90) | 3.51 (2.10) | 4.52 | .04* |
| Rate<sup>c</sup> | 145.14 (71.40) | 99.91 (77.30) | 4.62 | .04* |
| Duration<sup>d</sup> | 29.65 (9.29) | 23.41 (10.05) | 5.20 | .03* |

Note: \* $p < .05$ ; \*\* $p < .01$
<sup>a</sup>Proportion of hospital days with family visitation during study period.
<sup>b</sup>Frequency of DC instances per day during study period.
<sup>c</sup>Rate of DC expressed as minutes per day during study period.
<sup>d</sup>Duration of each instance of DC during study period.

Finally, we explored differences in the frequency, rate, and duration of DC activities delivered by *Family* versus *Clinical Staff*. Figure 1A shows the estimated marginal means for the number of instances of DC delivered by Family versus Clinical Staff for infants in the 2019 versus 2020 cohorts. A significant main effect of source, *F*(1,49)=7.2, *p* = 0.01, reflected the fact that Clinical Staff delivered more instances of DC than family overall, and a main effect of Cohort, *F*(1,49) = 4.5, *p* = 0.04, indicated that more instances of DC occurred for infants in the 2019 cohort overall compared to those in the 2020 cohort. Importantly, the Cohort by Source interaction was not statistically significant, *F*(1,49) = 0.01, *p* = 0.96, suggesting that the reduction in the number of instances of DC was the same regardless of whether Family or Clinical Staff were delivering the care.

**Figure 1.**
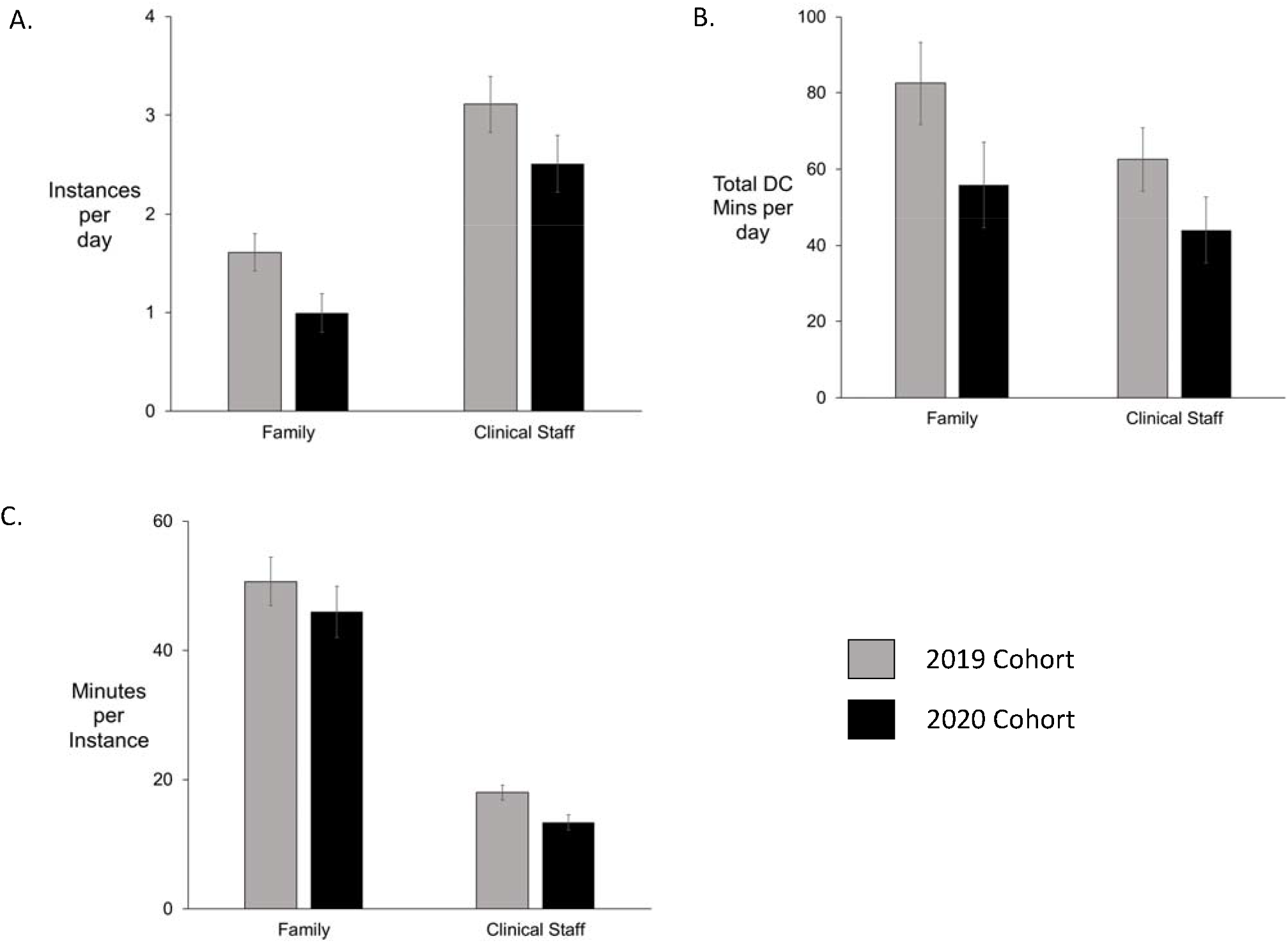
Estimated marginal means (SE) for DC frequency, amount, and duration by cohort (2019, 2020) and source (Family, Clinical Staff), controlling for PMA at start of study period (*N*=52).

Figure 1B shows a similar pattern for DC minutes per day. Even though family members were likely to deliver fewer instances of DC than clinical staff, family members contributed more to the overall amount of DC that infants experienced, *F*(1,49)=11.7, *p* = 0.001. As above, infants in the 2019 cohort experienced significantly more DC than infants in the 2020 cohort, F(1,49)=4.6, p = 0.04, yet a non-significant interaction indicated that the reduction between 2019 and 2020 was similar for both family- and clinical staff-delivered DC, *F*(1,49)=0.19, *p* = 0.67.

Figure 1C shows the mean duration of each instance of DC delivered by Family versus Clinical Staff. Here, DC delivered by family was significantly longer per instance than that delivered by clinical staff, *F*(1,49)=18.3, *p* < 0.001. However, the main effect of cohort, *F*(1,49)=2.2, *p* = 0.15, and the cohort by delivery source interaction, *F*(1,49)=0.01, *p* = 0.99, did not achieve statistical significance. This finding suggests that, although the impact of the COVID-19 pandemic-related hospital policies on DC were strongly reflected in the frequency and thereby, the overall amount of DC that infants received, the impact was less pronounced on the length of time that family and clinical staff spent delivering DC once that activity was undertaken.

## Discussion

Our study documents lower rates of family visitation, fewer instances of DC activities, by both family members and clinical staff, and shorter periods of DC on average for preterm infants cared for in the NICU during a COVID-19 pandemic-affected period in 2020 compared to a similar period in 2019. Parents in the COVID-19 period visited their infants roughly half as much as in the preceding year. Both frequency and length of DC activities were reduced whether provided by family or clinical staff. However, frequency was more impacted than length in our comparison. Surprisingly, a similar reduction in DC activities occurred if provided by clinical staff to that provided by family members. Although bedside nurses were unaffected by limitations to visitation, they were not immune to more subtle psychological impacts of a pandemic and may have felt undersupported by other staff members.

Concerns about the impacts of the COVID-19 pandemic on family visitation in the NICU and engagement have been raised^5,6^; however, our study is the first to document the concrete existence and degree of the feared impact. Although reduction in family visitation is not an unexpected finding considering the changes to hospital policies, it represents a significant threat to parent-infant bonding, the delivery of important parent-delivered care activities^9–11^, and positive health outcomes for parents and infants^12–14^. DC has shown to produce positive impacts on both short-term (cardiorespiratory stability, growth, infection rates) and long-term (neurodevelopmental) outcomes^15^, raising concerns that care may have been compromised in the NICU and that this impact may be long-lasting. Concerning also is the impact on parental mental health. Qualitative analysis of NICU parent surveys around COVID-19-related visitation restrictions revealed that over 50% of parents were experiencing increased sadness or anger and 25% were distressed by feelings of separation from either partners or the newborn^16^. It is somewhat heartening to see that frequency was more affected than duration; if parents were able to visit and engage with their infants, the length of engagement was only partially affected. Although minimum effective doses of these activities are still being explored, by reducing amounts of DC activities, possible decreases in the benefits may ensue. Close tracking of disturbances in family bonding, family mental health, and infant neurodevelopmental outcomes is warranted.

In addition to differences in family visitation and involvement in care, we also found differences in DC activities delivered by clinical staff, who should be unaffected by restrictions in visitation policies. Fear of transmission of infection may impact clinical staff behavior, reducing contact^17^ and even leading to an increase in medical errors. Similar concerns were raised during the Ebola outbreak^18^, where nurses felt the need for personal protective equipment (PPE) and the risks associated with touch negatively impacted the quality of their care. The importance of human touch for human wellbeing, both mental and physical, has been well described^19^. Concerns have been raised about touch starvation during the COVID-19 pandemic, particularly in adults for whom physical distancing may limit interpersonal contact^20^. Our study presents data suggesting reduced touch is occurring even in the NICU among infants who are at high risk of poor health outcomes.

Mitigation strategies should be implemented immediately. Recommendations for supporting parent-infant attachment during the COVID-19 pandemic have been reported^21^, many of which our institution was following during the 2020 period, with significant impacts measured on visitation and care despite best efforts. Alteration of social factors that may contribute to reduced visitation is difficult; lack of childcare, job losses, and fear of exposure outside the home cannot be changed by hospital policy. However, clinical staff should reduce barriers for family engagement when visitation does occur so that time may be of maximum benefit. Clinical staff may also attempt to consciously increase DC activities they provide to reduce, albeit only partially, the absence of the parent or other family members. Structured DC programs augmented by clinical staff when family members were not able to be present, showed health benefits for NICU infants^22^. Some opportunities may exist to support parent-infant attachment via technology^23^. Finally, government and hospital policymakers should carefully consider the possible negative impacts to patients and families when making decisions that may adversely affect immediate and long-term outcomes among NICU families. Although reduced visitation may decrease COVID-19 transmission risk to patients, families, and medical staff, it also impairs care that is both compassionate and medically indicated.

Limitations to this study include its reliance on bedside charting by nurses that may provide an incomplete picture of all interactions due to inadvertent omissions. When comparing two periods, one must also consider the effects of changing clinical staff. New nurses were hired between 2019 and 2020, but nurse educational onboarding included expectations for family visitation and DC charting. However, it is possible that the differences across study period that were documented here, at least partially, could relate to differences in nurse charting practices. Also limiting is the single-center nature of the study. Effects at our institution may or may not agree with those at other hospitals with different restrictions on family visitation or parent involvement in infant care. Some hospitals have forbidden all skin-to-skin care in their NICUs in an effort to reduce possible parent-infant COVID-19 transmission, while we have continued to allow it in asymptomatic parents who practice good hand hygiene and wear masks. Differences in visitation policies also exist across counties, cities, and states around the country, creating variation in effects. Other limitations include unavailable data on parent mood, infant health outcomes, and neurodevelopment. Also unexplored due to sample size was an evaluation of socioeconomic-related health care disparities, important to include in any outcome analysis. These areas remain good avenues for future research into the effects of this pandemic on at-risk infants and families who experience a NICU stay.

The COVID-19 pandemic has had a devasting impact on global health in ways both directly and indirectly measured. Although significant mortality and morbidity has been recorded particularly in older adult patients, infants have been largely spared^24^. However, indirect impacts of the COVID-19 pandemic on preterm infants in the NICU and their families are significant and, until now, largely unmeasured. Careful attention needs to be paid to current infant care patterns and hospital policies and to long-term follow up of infants in the NICU and their families.

## Data Availability

De-identified data are available upon request to senior author.

